## Supporting Information Table 1 for "COVID-19 isolation and quarantine orders in Berlin-Reinickendorf (Germany): How many, how long and to whom?"

**Supporting Information 1: Recommendations by the Robert Koch Institute on the duration of isolation, quarantine and contact person definition (original text excerpts)**

| | |
|---|---|
| **Isolation duration period 1 ($I_1$)** | |
| Time period: | 03.03.2020 to 01.07.2020 |
| Source: | Link to recommendations of the Robert Koch Institute on the Waybackmachine archive |
| Text extract: | Kriterien zur Entlassung aus der häuslichen Isolierung: Frühestens 14 Tage nach Symptombeginn UND Symptomfreiheit seit mind. 48 Stunden bezogen auf die akute COVID-19-Erkrankung |
| **Isolation duration period 2 ($I_2$)** | |
| Time period: | 02.07.2020 to 30.03.2021 |
| Source: | Link to recommendations of the Robert Koch Institute on the Waybackmachine archive |
| Text extract: | Mind. 48 Stunden Symptomfreiheit PLUS Frühestens 10 Tage nach Symptombeginn |
| **Isolation duration period 3 ($I_3$)** | |
| Time period: | 31.03.2021 to 18.12.2021 |
| Source: | Link to recommendations of the Robert Koch Institute on the Waybackmachine archive |
| Text extract: | Für Patienten mit leichtem oder mildem/ moderatem Krankheitsverlauf (gemäß WHO-Definition) und ungestörter Immunkompetenz kann eine Entisolierung erfolgen, wenn (1) mindestens 14 Tage seit Auftreten der ersten Symptome verstrichen sind, (2) eine nachhaltige Besserung der akuten COVID-19-Symptomatik gemäß ärztlicher Beurteilung seit >48 h vorliegt und (3) ein negativer Antigentest. |
| **Quarantine duration period 1 ($Q_1$)** | |
| Time period: | 03.03.2020 to 30.11.2020 |
| Source: | Link to recommendations of the Robert Koch Institute on the Waybackmachine archive |
| Text extract: | Gesundheitsüberwachung bis zum 14. Tag nach dem letzten Kontakt mit dem bestätigten 2019-nCoV-Fall. Kontaktreduktion nach Maßgabe des Gesundheitsamtes. Dies kann, gemäß § 31 IfSG, die Isolation im Krankenhaus beinhalten, oder eine Selbstisolierung während der weiteren diagnostischen Abklärung unter Einhaltung infektionshygienischer Maßnahmen. |
| **Quarantine duration period 2 ($Q_2$)** | |
| Time period: | 01.12.2020 to 15.02.2021 |
| Source: | Link to recommendations of the Robert Koch Institute on the Waybackmachine archive |
| Text extract: | Häusliche Absonderung für 14 Tage (Quarantäne) - gerechnet ab dem letzten Tag des Kontaktes zum Quellfall. Die häusliche Absonderung kann auf 10 Tage verkürzt werden, wenn ein negativer SARS-CoV-2-Test vorliegt; der Test darf frühestens am zehnten Tag der Quarantäne durchgeführt werden. |
| **Quarantine duration period 3 ($Q_3$)** | |
| Time period: | 16.02.2021 to 08.09.2021 |
| Source: | Link to recommendations of the Robert Koch Institute on the Waybackmachine archive |
| Text extract: | Kontaktpersonen der Kategorie 1 müssen sich unverzüglich für 14 Tage häuslich absondern (Quarantäne) - gerechnet ab dem letzten Tag des Kontaktes zum Quellfall. |
| **Quarantine duration period 4 ($Q_4$)** | |
| Time period: | 09.09.2021 to 18.12.2021 |
| Source: | Link to recommendations of the Robert Koch Institute on the Waybackmachine archive |
| Text extract: | 10 Tage Quarantäne ohne abschließenden Test. 5 Tage mit PCR-Test bei Probenentnahme frühestens am 5. Tag. Bei Personen, die regelmäßig im Rahmen einer seriellen Teststrategie getestet werden (z.B. Schülerinnen und Schüler), kann der negative Nachweis auch mittels qualitativ hochwertigen Antigen-Schnelltests erwogen werden. 7 Tage mit Antigen-Schnelltest bei Probenentnahme frühestens am 7. Tag. |
| **Contact person definition period 1 ($C_1$)** | |
| Time period: | 03.03.2020 to 30.03.2021 |
| Source: | Link to recommendations of the Robert Koch Institute on the Waybackmachine archive |
| Text extract: | Personen mit kumulativ mindestens 15-minütigem Gesichts- (face-to-face) Kontakt, z.B. im Rahmen eines Gesprächs. Dazu gehören z.B. Personen aus Lebensgemeinschaften im selben Haushalt. Personen mit direktem Kontakt zu Sekreten oder Körperflüssigkeiten, insbesondere zu respiratorischen Sekreten eines bestätigten COVID-19-Falls, wie z.B. Küssen, Kontakt zu Erbrochenem, Mund-zu-Mund Beatmung, Anhusten, Anniesen, etc. Personen, die nach Risikobewertung durch das Gesundheitsamt mit hoher Wahrscheinlichkeit einer relevanten Konzentration von Aerosolen ausgesetzt waren (z.B. Feiern, gemeinsames Singen oder Sporttreiben in Innenräumen) Medizinisches Personal mit Kontakt zum bestätigten COVID-19-Fall im Rahmen von Pflege oder medizinischer Untersuchung (<= 2m), ohne verwendete Schutzausrüstung. Falls die Person früher als COVID-19 Fall gemeldet wurde ist keine Quarantäne erforderlich, es soll ein Selbstmonitoring erfolgen und bei Auftreten von Symptomen eine sofortige Selbst-Isolation und -Testung. Bei positivem Test wird die Kontaktperson zu einem Fall. Bei diesem sollten alle Maßnahmen ergriffen werden wie bei sonstigen Fällen auch (inkl. Isolation). Kontaktpersonen eines bestätigten COVID-19-Falls im Flugzeug: Passagiere, die direkter Sitznachbar des bestätigten COVID-19-Falls waren, unabhängig von der Flugzeit. Saß der COVID-19-Fall am Gang, so zählt der Passagier in derselben Reihe jenseits des Ganges nicht als Kontaktperson der Kategorie I, sondern als Kontaktperson der Kategorie II. Besatzungsmitglieder oder andere Passagiere, sofern auf Hinweis des bestätigten COVID-19-Falls eines der anderen Kriterien zutrifft (z.B. längeres Gespräch; o.ä.). |
| **Contact person definition period 2 ($C_2$)** | |
| Time period: | 31.03.2021 to 19.05.2021 |
| Source: | Link to recommendations of the Robert Koch Institute on the Waybackmachine archive |
| Text extract: | Enger Kontakt <1,5 m, Nahfeld) länger als 10 Minuten ohne adäquaten Schutz (adäquater Schutz = Fall und Kontaktperson tragen durchgehend und korrekt MNS [Mund-Nasen-Schutz] oder FFP2-Maske). Gespräch mit dem Fall (face-to-face-Kontakt, <1,5 m, unabhängig von der Gesprächsdauer) ohne adäquaten Schutz (adäquater Schutz = Fall und Kontaktperson tragen durchgehend und korrekt MNS [Mund-Nasen-Schutz] oder FFP2-Maske). Gleichzeitiger Aufenthalt von Kontaktperson und Fall im selben Raum mit wahrscheinlich hoher Konzentration infektiöser Aerosole unabhängig vom Abstand für >10 Minuten, auch wenn durchgehend und korrekt MNS (Mund-Nasen-Schutz) oder FFP2-Maske getragen wurde. Vollständig gegen COVID-19 geimpfte Personen sind nach Exposition zu einem bestätigten SARS-CoV-2-Fall von Quarantäne-Maßnahmen ausgenommen, ebenso wie Personen, die in der Vergangenheit eine PCR-bestätigte und symptomatische COVID-19-Erkrankung durchgemacht haben ("Genesene") und mit einer Impfstoffdosis geimpft sind. |
| **Contact person definition period 3 ($C_3$)** | |
| Time period: | 20.05.2021 to 18.12.2021 |
| Source: | Internal guideline document by the local public health agency |
| Text extract: | Enger Kontakt (<1,5 m, Nahfeld) länger als 10 Minuten ohne adäquaten Schutz (adäquater Schutz = Fall und Kontaktperson tragen durchgehend und korrekt MNS (Mund-Nasen-Schutz) oder FFP2-Maske). Gespräch mit dem Fall (Face-to-face-Kontakt, <1,5 m, unabhängig von der Gesprächsdauer) ohne adäquaten Schutz (adäquater Schutz = Fall und Kontaktperson tragen durchgehend und korrekt MNS (Mund-Nasen-Schutz) oder FFP2-Maske) oder direkter Kontakt (mit respiratorischem Sekret). Gleichzeitiger Aufenthalt von Kontaktperson und Fall im selben Raum mit wahrscheinlich hoher Konzentration infektiöser Aerosole unabhängig vom Abstand für >10 Minuten, auch wenn durchgehend und korrekt MNS (Mund-Nasen-Schutz) oder FFP2-Maske getragen wurde.<br>Coronavirus-Update (Internes Dokument des Gesundheitsamtes Reinickendorf): Umgang mit Kontaktpersonen in Schulen Wenn wir einen Fall in einer Schulklasse hatten, wird die gesamte Klasse über eine Fernfeldübertragung nur noch bei den folgenden Konstellationen in Quarantäne gesteckt: 1) Es sind schon Übertragungen in der Klasse in der zu bewertenden Situation bekannt geworden 2) Es wurde gar nicht gelüftet und die Zeit zusammen im Raum war über 2h. 3) Sonderfälle, bei dem grob viele Hygieneregeln missachtet wurden, nach Rücksprache. Davon unbenommen bleibt aber, dass die Personen durch eine Nahfeldübertragung Kontaktpersonen geworden sind. Zum Beispiel die besten Kumpels in der Klasse, die eng sitzenden Sitznachbarn, die Erziehungskraft, die das Kind ganz eng betreut hat. |
