## Supporting Information Figure 2 for "COVID-19 isolation and quarantine orders in Berlin-Reinickendorf (Germany): How many, how long and to whom?"

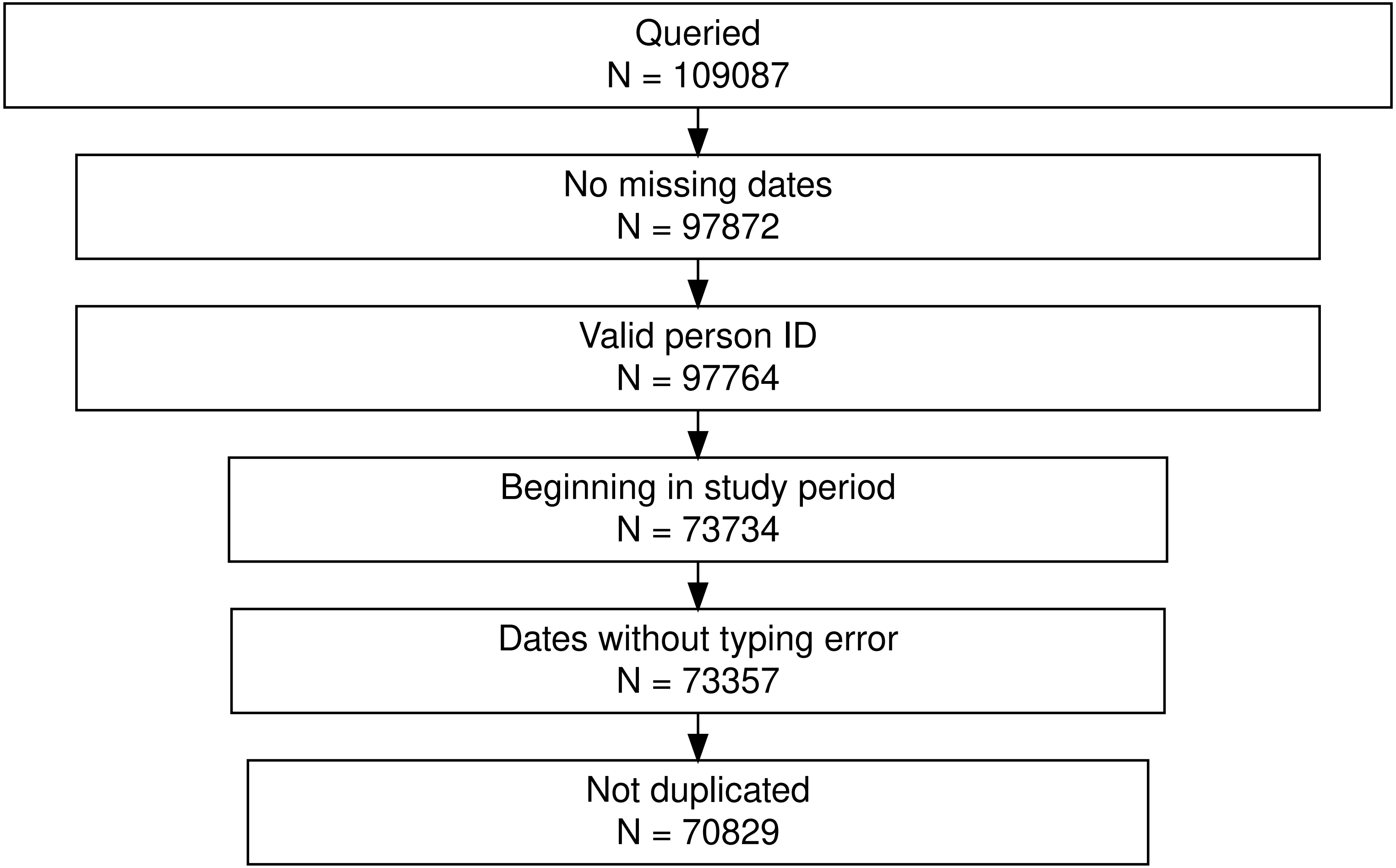


**Supporting Information 2: Inclusion and exclusion criteria for the data cleaning process** from database export (queried) to data entries that were included in the final analysis (not duplicated).
